# Delirium is associated with incident dementia across the multimorbidity spectrum: a population-based cohort study

**DOI:** 10.1101/2025.10.06.25337244

**Authors:** Rose S Penfold, Clare MacRae, Elizabeth L Sampson, Daniel HJ Davis, Atul Anand, E. Wesley Ely, Bruce Guthrie, Alasdair MJ MacLullich

**Affiliations:** Usher Institute, University of Edinburgh, 5-7 Little France Rd, Edinburgh, EH16 4UX; Advanced Care Research Centre, University of Edinburgh, Edinburgh, EH16 4UX; Academic Centre for Healthy Ageing, Whipps Cross Hospital, Barts NHS Trust, E11 1NR & Centre for Psychiatry and Mental Health, Wolfson Institute of Population Health, Queen Mary University of London, London, E13 8SP; Centre for Cardiovascular Sciences, Queen’s Medical Research Institute, University of Edinburgh, EH16 4TJ; MRC Unit for Lifelong Health and Ageing, University College London, WC1E 7HB; The Critical Illness, Brain Dysfunction, and Survivorship (CIBS) Center, Vanderbilt University Medical Center, & The Veterans Affairs Tennessee Valley Geriatric Research Education and Clinical Center (GRECC), Nashville TN

**Author notes:** **Corresponding author** Rose S Penfold, Usher Institute, Usher Building, The University of Edinburgh, 5-7 Little France Rd, Edinburgh, EH16 4UX. joint last authorship (Bruce Guthrie, Alasdair MJ MacLullich).

## Abstract

**Background:** Delirium is strongly associated with subsequent dementia, but this is often assumed to reflect underlying associations of baseline health with dementia. We examined the associations of delirium on admission with incident dementia across the spectrum of multimorbidity.

**Methods:** Retrospective population-based cohort study using linked primary care and hospital data for emergency admissions aged ≥65 years in Lothian, Scotland, from 1 Apr 2017 to 1 Apr 2020. Delirium on admission was assessed at the bedside for all patients using the 4AT (www.the4AT.com). Associations of delirium, multimorbidity, and their interaction with incident dementia and mortality were examined using Fine-Gray competing-risks regression and Cox proportional hazards models.

**Results:** Of 23,558 people without pre-existing dementia (mean age 78.9 years, 54% female), 4135 (18%) had an admission with delirium. Delirium was associated with higher incident dementia risk. The relative risk was highest in those without multimorbidity (adjusted Hazard Ratio (aHR) 3.38, 95% CI 2.46–4.63) and decreased with an increasing number of long-term conditions. Delirium was also associated with increased mortality, regardless of multimorbidity. In those without multimorbidity, delirium was associated with higher early mortality (≤90 days: aHR 4.23, 95% CI 3.27 to 5.49) and late mortality (>90 days: aHR 1.64, 95% CI 1.33 to 2.03).

**Conclusion:** Delirium is strongly associated with incident dementia in older adults across the multimorbidity spectrum, with the highest relative risk in people without multimorbidity. Findings support routine delirium assessment on hospital admission for all older adults and highlight need to further investigate neurodegenerative mechanisms in delirium.

## Introduction

Delirium is an acute neuropsychiatric condition with multiple potential triggers, affecting 23% of acutely hospitalised older adults,^1^ or around 1.8 million in the UK and over 7 million in the USA each year.^2,3^ It is associated with higher mortality, longer hospital stays, and higher care home admission rates.^4^ Existing evidence strongly supports a link between delirium and subsequent dementia. A 2020 meta-analysis of 23 studies (n=10,369) reported an association between delirium and long-term cognitive decline across medical and surgical cohorts (estimated effect size Hedges’ *g* = 0.45; odds ratio ≈ 2.3).^5^ Subsequent studies support these findings.^6,7^ The DECIDE study demonstrated that the association between delirium and dementia persists after adjusting for baseline cognition and illness severity,^6^ and a 2024 large hospital data study reported a dose-dependent association between number of delirium episodes and incident dementia, supporting a causal interpretation.^7^

Multimorbidity is commonly defined as the co-existence of two or more long-term conditions (LTC).^8^ Multimorbidity is a risk factor for delirium,^9^ and, like delirium, is associated with longer hospital stays, higher healthcare costs, higher mortality,^10^ and subsequent dementia.^11-13^A 2024 scoping review identified 140 studies examining delirium and multimorbidity in combination,^9^ but only six explored delirium outcomes, and just one focused on cognitive decline.^14^ That study, in a subgroup of 126 older elective surgical patients without dementia, found that multimorbidity explained some observed variation in cognitive decline following delirium. Another study in 821 critically ill patients demonstrated that delirium was associated with cognitive decline at one year, even in younger adults without preexisting conditions.^15^ However, it remains unclear whether the association between delirium and future dementia risk reflects an increased vulnerability to delirium in people with multimorbidity, or an independent association of delirium. Additionally, it is unknown whether multimorbidity modifies the relationship between delirium and dementia. This is of clinical importance, because an independent association suggests effective delirium prevention or treatment could help reduce future dementia risk.

The aim of this study was to investigate the associations of delirium and multimorbidity with incident dementia in a population-based cohort of acutely hospitalised older adults. A secondary aim was to investigate associations with mortality.

## Methods

### Study design and population

This retrospective population-based cohort study used routinely collected pseudonymised data accessed via the DataLoch service (dataloch.org). Information on emergency hospitalisations was obtained from the hospital electronic health records (EHR) system (TrakCare, Intersystems), capturing all admissions to three acute hospitals in NHS Lothian (a large health board in Southeast Scotland with a population ∼ 900,000). The study cohort was drawn from a dataset which was available to investigators and includes all adults registered with a General Practitioner (GP) in NHS Lothian contributing to DataLoch (∼90% of GP practices and ∼86% of older adults in the health board), with at least one year of GP registration.^16^

### Study cohort

The processes for cohort selection are shown in Supplementary Figure 1. A three-year study period was defined (1 April 2017 to 1 April 2020) to ensure availability of 4AT delirium assessment data in hospital EHR and sufficient follow-up after admission. All patients had at least 42 months (3.5 years) follow-up from the end of the study period until study end (23 October 2023).

We identified all emergency medical and orthopaedic admissions during the study period for people aged ≥65 years at admission within the eligible study population. We categorised people with any eligible admission into two mutually exclusive groups: those with one or more admissions with a 4AT score ≥4 during the study period (delirium group), and those with one or more admissions with 4AT scores between 0-3, and no score ≥4 (no delirium group). In the delirium group, study entry was defined as the date of the first admission with delirium (index admission). In the no delirium group, study entry was defined as the date of the first admission with a recorded 4AT score (index admission). Number of days until study end was similar between the two groups (Supplementary Table 1).

We excluded individuals who had a dementia diagnosis prior to or during their index admission, as incident dementia was a primary outcome, and those without a recorded 4AT score during the study period.

### Outcomes

Information on dementia diagnoses was obtained from primary care, hospital discharge, and community prescribing records linked via a unique individual identifier. Time to incident dementia diagnosis was determined by the date of first diagnosis appearing in any data source, with dementia ascertained in GP and hospital discharge data using code lists from the Health Data Research (HDR) UK phenotype library (Supplementary Table 2). Information on community-dispensed prescriptions was obtained through linkage to the national Prescribing Information System (PIS). In Scotland, all prescribed medications are provided by the National Health Service (NHS) based on clinical need without additional cost to individuals. Donepezil, galantamine, rivastigmine, and memantine are only recommended for NHS prescription by the Scottish Medicines Consortium for dementia treatment, and prescription of any one was considered diagnostic of dementia. Mortality was determined from National Records of Scotland (NRS) mortality registration data, capturing all deaths in Scotland.

### Covariates

#### Delirium

Delirium assessment is embedded as part of routine care on admission for all medical and orthopaedic admissions aged ≥65 years to Lothian hospitals using the 4AT (www.the4AT.com), an extensively validated tool with good sensitivity and specificity recommended in UK national guidelines.^17,18^ Recording of 4AT scores in hospital EHR systems has been routine since April 2016. We have previously demonstrated high 4AT completion rates for medical (∼80%) and orthopaedic (>90%) admissions in this population.^19,20^

#### Multimorbidity

We used a list of LTC recommended for inclusion in multimorbidity research by an international Delphi consensus study,^21^ with LTC ascertained using code lists from the HDR UK phenotype library (Supplementary Table 3).

Participants were defined as having a LTC if a relevant code appeared in their primary care and/or hospital discharge records at any time before or during the index admission, except for cancers where lookback was limited to the preceding five years. Primary care data were obtained by linkage to GP records (Read version 2 codes), and hospital discharge data through linkage to Scottish Morbidity Records 01 (SMR01), a national administrative dataset capturing all acute inpatient care episodes, with one primary and up to five secondary diagnoses recorded for each episode using the International Classification of Diseases 10^th^ revision (ICD-10).

Individuals were categorised into four groups: no multimorbidity (0-1 LTC), 2-4 LTC, 5-6 LTC, and 7+ LTC, where the three categories of people with multimorbidity represent tertiles of number of LTC in the study cohort (Supplementary Figure 2).

#### Other variables

The Scottish Index of Multiple Deprivation (SIMD) is a small area measure of relative deprivation commonly used in health research.^22^ Information on sex and ethnicity were obtained from GP records, as recorded at registration.

#### Analyses

All analyses were performed using de-identified data within a secure data environment approved by DataLoch, using R version 4.5.1. All categorical variables are presented as frequencies and percentages and continuous variables as mean (standard deviation) or median (interquartile range).

A cumulative incidence function (CIF) was plotted to visualise risk of incident dementia, accounting for competing mortality risk, and a Kaplan-Meier curve to visualise risk of death.

The key exposures were delirium, number of LTC, and their interaction, examined based on prior hypothesis and existing evidence suggesting that an individual’s baseline health may influence their vulnerability to the effects of delirium. Fine-Gray subdistribution hazards models were used to estimate the associations of key exposures with incident dementia, accounting for competing mortality risk. Model 1 included number of LTC, delirium, and their interaction. Model 2 was additionally adjusted for age, sex, and SIMD. Model fit was assessed using the Akaike Information Criterion (AIC) to compare models with and without the interaction term, and likelihood ratio tests to assess whether inclusion of interaction terms improved fit. Follow-up was from date of hospitalisation until first recorded dementia diagnosis, accounting for competing mortality risk, with those who remained event free censored on 23 October 2023.

Cox proportional hazards models were used to estimate the associations of key exposures with mortality. Two models were analysed, using the same modelling strategy described above. The proportional hazards assumption was tested using the ‘cox.zph’ function in R and visual inspection of Schoenfeld residuals. Since the proportional hazards assumption was violated for the delirium and LTC variables, a piecewise model was applied to estimate hazard ratios within two predefined time intervals: **≤**90 days and >90 days.^23^ These intervals were chosen based on clinical knowledge and inspection of the Kaplan-Meier curve, with 90 days selected to distinguish early from late mortality. Follow-up was from hospitalisation date until death, with those who remained event free censored on 23 October 2023.

A sensitivity analysis was performed with dementia as the outcome to assess the potential impact of undiagnosed dementia on the results. This excluded patients who died or were diagnosed with dementia within six months of their index admission.

#### Ethics

The project received approval through DataLoch, including: Caldicott Guardian approval, ACCORD sponsorship (AC23107), favourable ethical opinion under DataLoch’s delegated authority (Reference: 22/NS/0093), and a public value assessment by the DataLoch Public Reference Group.

## Results

### Baseline Characteristics

There were 58,290 emergency medical and orthopaedic admissions during the study period, involving 31,834 unique individuals (Supplementary Table 4). Of these, 5065 (15.9%) had a recorded dementia diagnosis, prior to admission (4442; 14.0%) or during their index admission (623; 2.0%), and were excluded. A further 3211 people without a recorded 4AT score during the study period were excluded. Characteristics of excluded individuals are in Supplementary Table 5.

Table 1 shows characteristics of the study cohort, stratified by delirium status. A total of 4135 (17.6%) people had at least one admission with delirium during the study period. Compared to those without delirium, people with delirium were on average older (mean age 81.9 vs. 78.3 years, p<0.001), more frequently female (55.6% vs. 54.2%, p<0.001), had a higher number of LTC (median 6 vs. 5 conditions, p<0.001), and longer hospital stays (median 9 vs. 4 days, p<0.001).

**Table 1:**
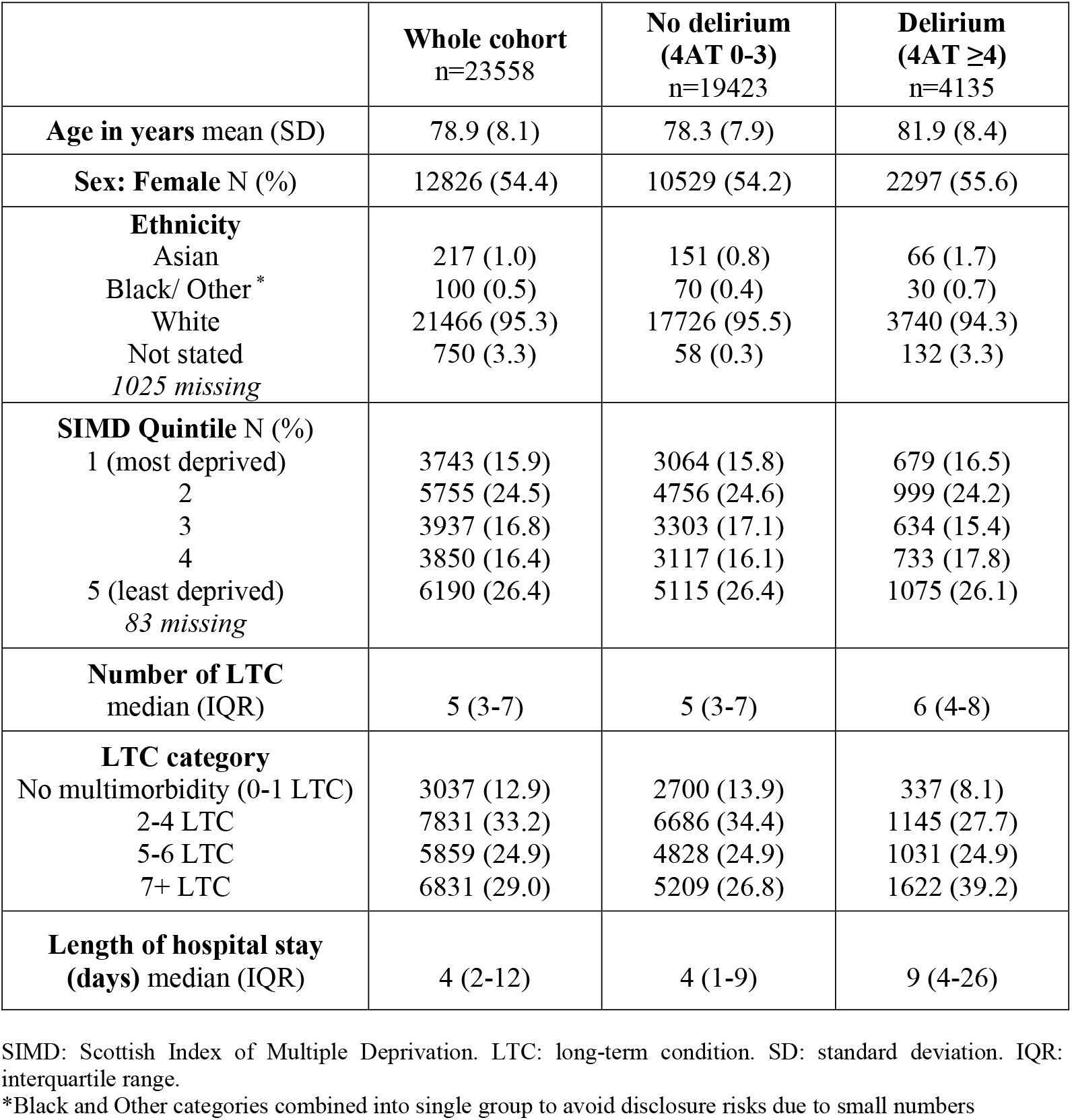
Characteristics of the study cohort at index admission, by delirium status.

|  | **Whole cohort**<br>n=23558 | **No delirium**<br>**(4AT 0-3)**<br>n=19423 | **Delirium**<br>**(4AT ≥4)**<br>n=4135 |
| --- | --- | --- | --- |
| **Age in years** mean (SD) | 78.9 (8.1) | 78.3 (7.9) | 81.9 (8.4) |
| **Sex: Female** N (%) | 12826 (54.4) | 10529 (54.2) | 2297 (55.6) |
| **Ethnicity**<br>Asian<br>Black/ Other *<br>White<br>Not stated<br>*1025 missing* | 217 (1.0)<br>100 (0.5)<br>21466 (95.3)<br>750 (3.3) | 151 (0.8)<br>70 (0.4)<br>17726 (95.5)<br>58 (0.3) | 66 (1.7)<br>30 (0.7)<br>3740 (94.3)<br>132 (3.3) |
| **SIMD Quintile** N (%)<br>1 (most deprived)<br>2<br>3<br>4<br>5 (least deprived)<br>*83 missing* | 3743 (15.9)<br>5755 (24.5)<br>3937 (16.8)<br>3850 (16.4)<br>6190 (26.4) | 3064 (15.8)<br>4756 (24.6)<br>3303 (17.1)<br>3117 (16.1)<br>5115 (26.4) | 679 (16.5)<br>999 (24.2)<br>634 (15.4)<br>733 (17.8)<br>1075 (26.1) |
| **Number of LTC**<br>median (IQR) | 5 (3-7) | 5 (3-7) | 6 (4-8) |
| **LTC category**<br>No multimorbidity (0-1 LTC)<br>2-4 LTC<br>5-6 LTC<br>7+ LTC | 3037 (12.9)<br>7831 (33.2)<br>5859 (24.9)<br>6831 (29.0) | 2700 (13.9)<br>6686 (34.4)<br>4828 (24.9)<br>5209 (26.8) | 337 (8.1)<br>1145 (27.7)<br>1031 (24.9)<br>1622 (39.2) |
| **Length of hospital stay**<br>**(days)** median (IQR) | 4 (2-12) | 4 (1-9) | 9 (4-26) |

SIMD: Scottish Index of Multiple Deprivation. LTC: long-term condition. SD: standard deviation. IQR: interquartile range.
\*Black and Other categories combined into single group to avoid disclosure risks due to small numbers

### Associations of Delirium and Multimorbidity with Incident Dementia

Figure 1 shows the cumulative incidence of incident dementia, accounting for competing mortality risk, stratified by delirium status and number of LTC. The overall dementia diagnosis rate in the cohort was 3.3 per 100 person-years (py) (95% CI 3.2 to 3.5), with higher rates in people with delirium across all multimorbidity levels, and in people with multimorbidity (Supplementary Table 6).

**Figure 1:**
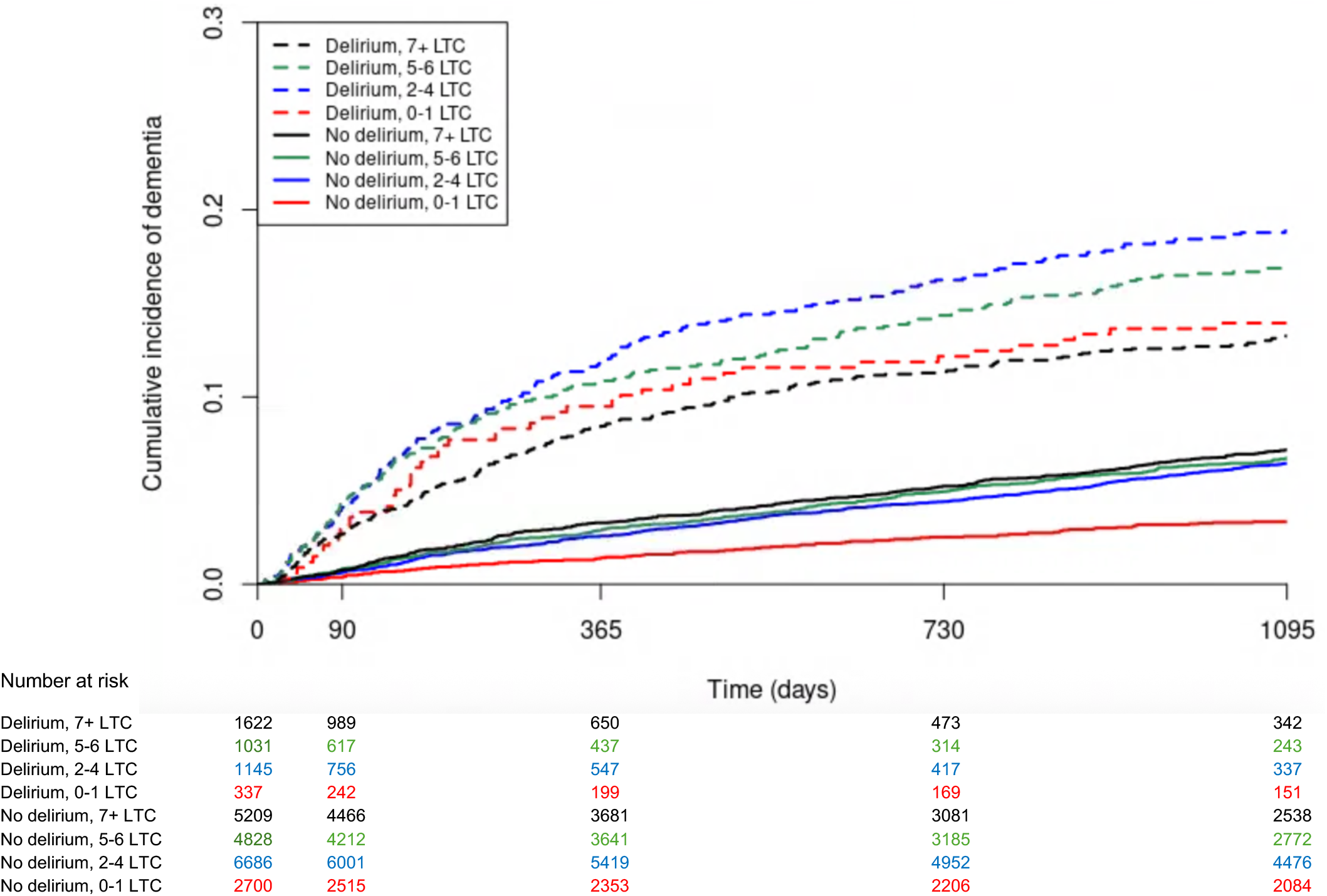
Cumulative incidence of a new dementia diagnosis accounting for competing risk of death for 23,558 adults ≥65 years with an emergency hospitalisation with a recorded 4AT score from 1 April 2017 to 1 April 2020, by delirium status and number of long-term conditions (LTC).

In people with delirium, those without multimorbidity had the highest relative risk of incident dementia, accounting for competing mortality risk (adjusted subdistribution HR 3.38, 95% CI 2.46 to 4.63; Table 2; Figure 3). Hazard ratios were progressively smaller with increasing number of LTC, although associations remained significant (for example, for people with delirium and 7+ LTC compared to people without delirium or multimorbidity, adjusted subdistribution HR 1.92, 95% CI 1.16 to 3.20). At one year following admission with delirium, 32/337 (9.5%) of people without multimorbidity had received a dementia diagnosis (Supplementary Table 7).

**Table 2:**
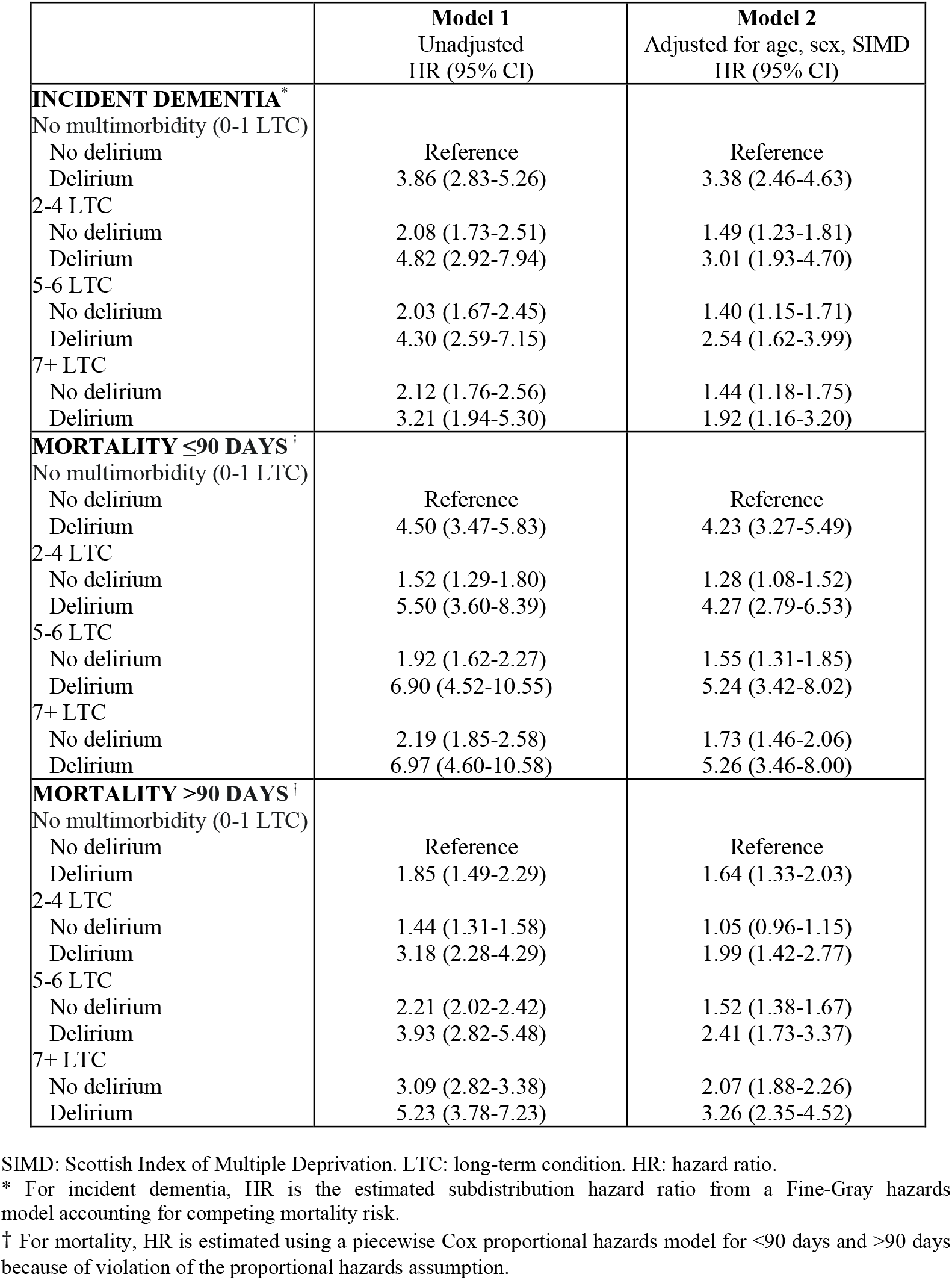
Unadjusted and adjusted subdistribution hazard ratios for incident dementia and hazard ratios for mortality in hospitalised older adults with and without delirium, and by number of long-term conditions (LTC).

|  | **Model 1**<br>Unadjusted<br>HR (95% CI) | **Model 2**<br>Adjusted for age, sex, SIMD<br>HR (95% CI) |
| --- | --- | --- |
| **INCIDENT DEMENTIA\*** |  |  |
| No multimorbidity (0-1 LTC) |  |  |
| No delirium | Reference | Reference |
| Delirium | 3.86 (2.83-5.26) | 3.38 (2.46-4.63) |
| 2-4 LTC |  |  |
| No delirium | 2.08 (1.73-2.51) | 1.49 (1.23-1.81) |
| Delirium | 4.82 (2.92-7.94) | 3.01 (1.93-4.70) |
| 5-6 LTC |  |  |
| No delirium | 2.03 (1.67-2.45) | 1.40 (1.15-1.71) |
| Delirium | 4.30 (2.59-7.15) | 2.54 (1.62-3.99) |
| 7+ LTC |  |  |
| No delirium | 2.12 (1.76-2.56) | 1.44 (1.18-1.75) |
| Delirium | 3.21 (1.94-5.30) | 1.92 (1.16-3.20) |
| **MORTALITY ≤90 DAYS†** |  |  |
| No multimorbidity (0-1 LTC) |  |  |
| No delirium | Reference | Reference |
| Delirium | 4.50 (3.47-5.83) | 4.23 (3.27-5.49) |
| 2-4 LTC |  |  |
| No delirium | 1.52 (1.29-1.80) | 1.28 (1.08-1.52) |
| Delirium | 5.50 (3.60-8.39) | 4.27 (2.79-6.53) |
| 5-6 LTC |  |  |
| No delirium | 1.92 (1.62-2.27) | 1.55 (1.31-1.85) |
| Delirium | 6.90 (4.52-10.55) | 5.24 (3.42-8.02) |
| 7+ LTC |  |  |
| No delirium | 2.19 (1.85-2.58) | 1.73 (1.46-2.06) |
| Delirium | 6.97 (4.60-10.58) | 5.26 (3.46-8.00) |
| **MORTALITY >90 DAYS†** |  |  |
| No multimorbidity (0-1 LTC) |  |  |
| No delirium | Reference | Reference |
| Delirium | 1.85 (1.49-2.29) | 1.64 (1.33-2.03) |
| 2-4 LTC |  |  |
| No delirium | 1.44 (1.31-1.58) | 1.05 (0.96-1.15) |
| Delirium | 3.18 (2.28-4.29) | 1.99 (1.42-2.77) |
| 5-6 LTC |  |  |
| No delirium | 2.21 (2.02-2.42) | 1.52 (1.38-1.67) |
| Delirium | 3.93 (2.82-5.48) | 2.41 (1.73-3.37) |
| 7+ LTC |  |  |
| No delirium | 3.09 (2.82-3.38) | 2.07 (1.88-2.26) |
| Delirium | 5.23 (3.78-7.23) | 3.26 (2.35-4.52) |

SIMD: Scottish Index of Multiple Deprivation. LTC: long-term condition. HR: hazard ratio.
\* For incident dementia, HR is the estimated subdistribution hazard ratio from a Fine-Gray hazards model accounting for competing mortality risk.
† For mortality, HR is estimated using a piecewise Cox proportional hazards model for ≤90 days and >90 days because of violation of the proportional hazards assumption.

In people without delirium, those with 2-4 LTC had a higher risk of incident dementia compared to those without multimorbidity (adjusted subdistribution HR 1.49, 95% CI 1.23 to 1.81). There was no further increase in hazard ratios with a higher number of LTC.

In a sensitivity analysis excluding people who died or developed dementia within six months of hospital discharge, associations between delirium and incident dementia were slightly stronger across all multimorbidity levels (Supplementary Table 8). In people without delirium, associations were consistent.

### Associations of Delirium and Multimorbidity with Mortality

The Kaplan-Meier-derived curves in Figure 2 demonstrate incidence of death, stratified by delirium and number of LTC. The mortality rate in the whole cohort was 10.3 deaths per 100py (95% CI 10.1 to 10.4), with higher rates observed in people with delirium and in those with more LTC (Supplementary Table 6). The highest mortality rate was in those with delirium and 7+ LTC (16.4 deaths/100 py, 95% CI 15.5 to 17.3), and the lowest in those without delirium and without multimorbidity (5.8/100py, 95% CI 5.4 to 6.2).

**Figure 2:**
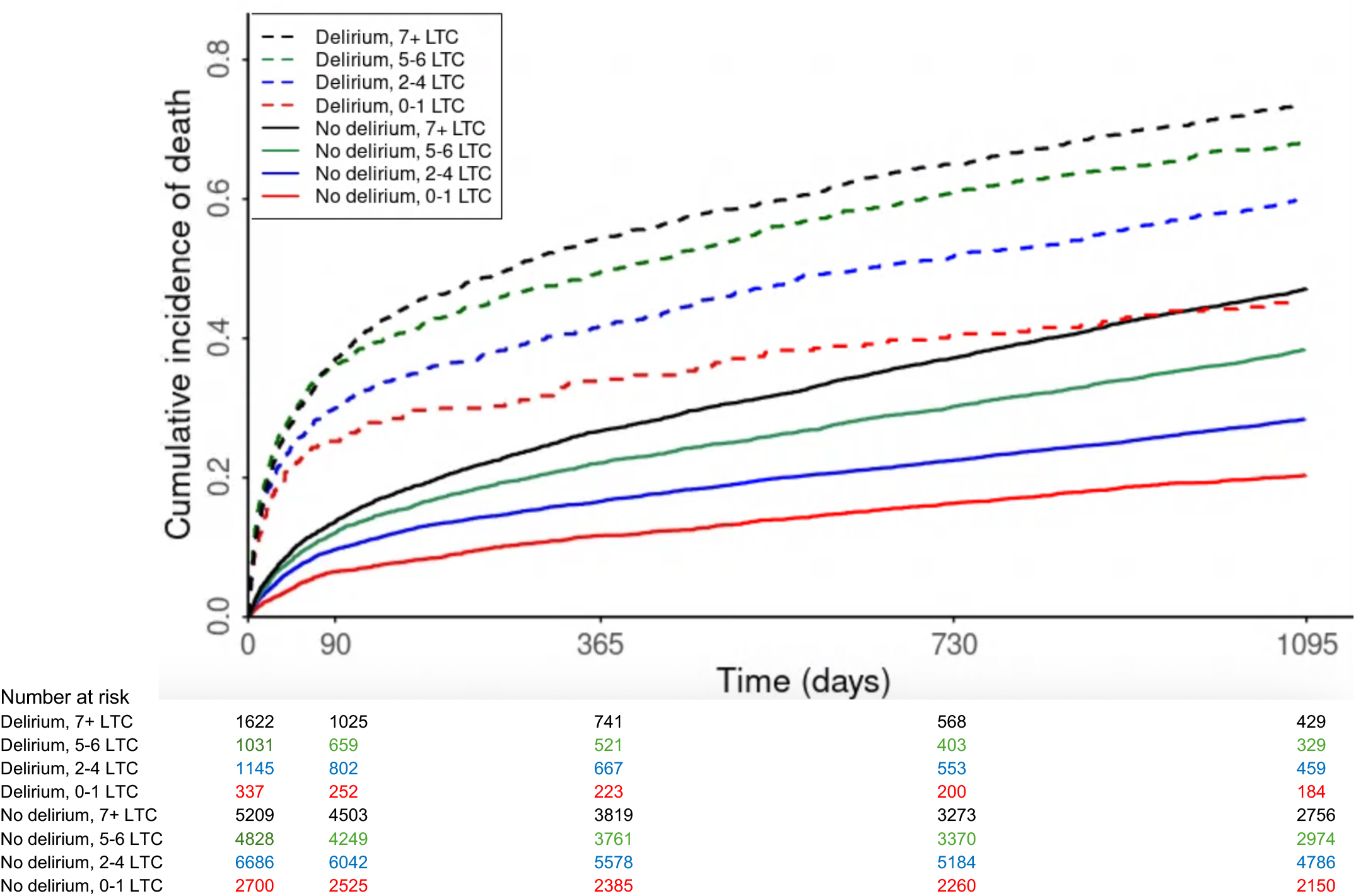
Kaplan-Meier estimates of cumulative mortality for 23,558 adults ≥65 years with an emergency hospitalisation with a recorded 4AT score from 1 April 2017 to 1 April 2020, by delirium status and number of long-term conditions (LTC).

Delirium was consistently associated with higher mortality risk, irrespective of the number of LTC. For example, in people without multimorbidity (0-1 LTC), delirium was associated with fourfold higher early mortality (≤90 days: adjusted Hazard Ratio (aHR) 4.23, 95% CI 3.27 to 5.49) and with higher late mortality (>90 days: aHR 1.64, 95% CI 1.33 to 2.03) in the adjusted model (Table 2; Figure 3). Early mortality (≤90 days) was significantly higher regardless of the number of LTC, but a higher number of LTC was associated with increasing late mortality (>90 days). At 90 days following admission with delirium, 85/337 (25.2%) of people without multimorbidity had died (Supplementary Table 7).

**Figure 3:**
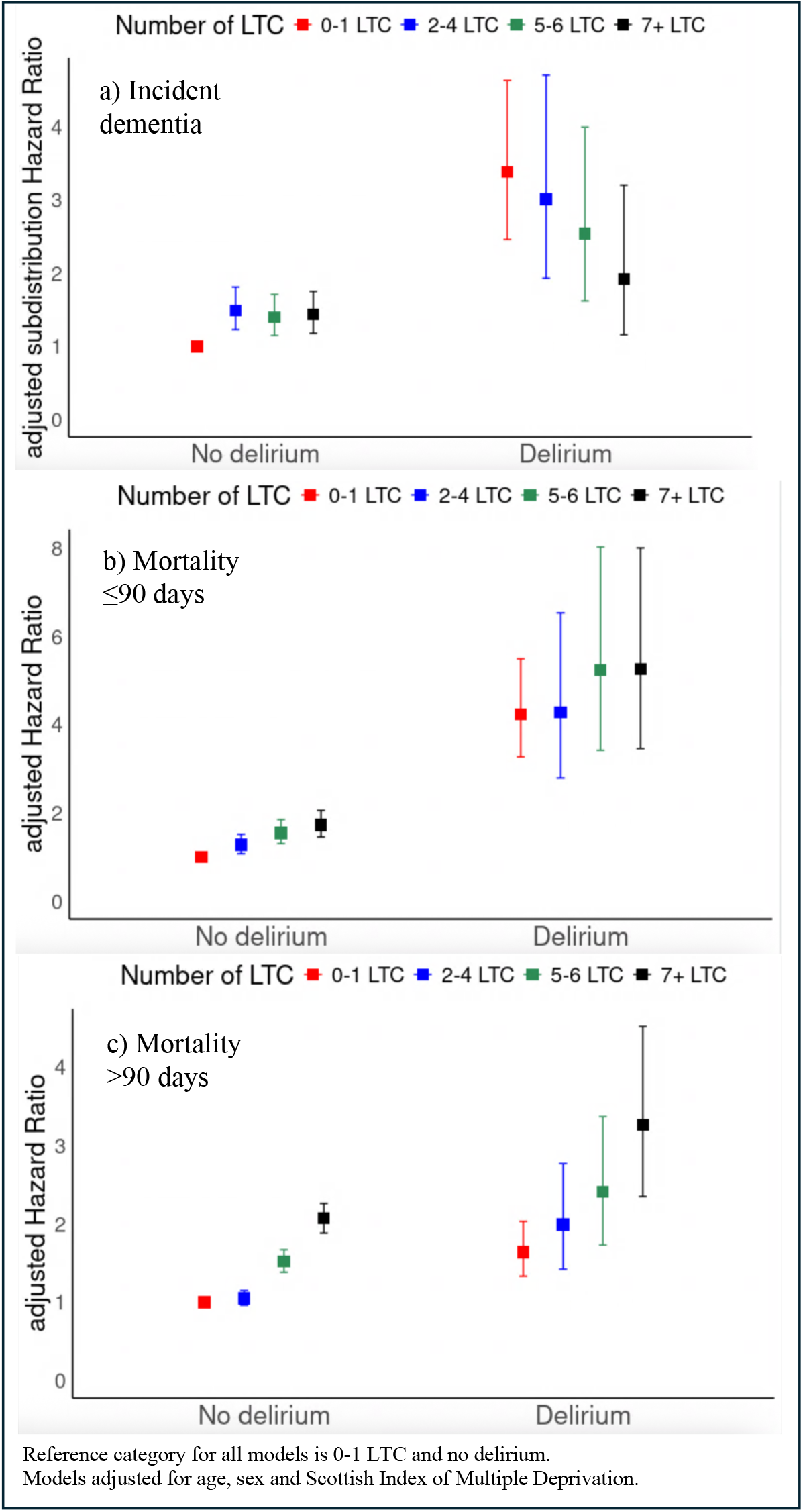
Adjusted subdistribution hazard ratios and 95% CIs for (a) incident dementia, and hazard ratios and 95% CIs for (b) mortality ≤90 days, and (c) mortality >90days, by delirium status and number of long-term conditions (LTC), based on interaction modelling.

In people without delirium, increasing number of LTC was associated with higher early (≤90 days) and late mortality (>90 days). For example, people with 7+ LTC had higher mortality compared to those without multimorbidity (≤90 days: aHR 1.73, 95% CI 1.46 to 2.06 and >90 days: aHR 2.07, 95% CI 1.88 to 2.26).

## Discussion

In this regional UK cohort of acutely hospitalised older adults, delirium was strongly associated with incident dementia across the multimorbidity spectrum. In people without multimorbidity, delirium was associated with threefold higher incident dementia risk, and relative risk decreased with an increasing number of long-term conditions (LTC). Delirium was also associated with higher mortality, regardless of number of LTC. At one year following an admission with delirium, 9.5% of people without multimorbidity had a new dementia diagnosis and 31.8% had died.

Taken together, although multimorbidity was more common in people with delirium, high incident dementia risk was not explained by multimorbidity and other individual characteristics. In fact, risk was relatively higher in people with delirium who did *not* have multimorbidity than those who did. These findings challenge any assumption that delirium is merely an epiphenomenon of baseline vulnerability, suggesting it is an important independent prognostic indicator. Delirium, and possibly other acute illness-related factors, may represent additional potentially modifiable contributors to dementia risk in hospitalised older adults beyond multimorbidity, complementing the Lancet Commission’s estimated ∼45% population attributable fraction from potentially modifiable risk factors.^11^

Previous meta-analyses have examined associations between delirium and dementia.^4,5^ However, many included studies used small, selected cohorts, variable methods for delirium and dementia ascertainment, and did not account for competing mortality risk. Effect estimates are therefore heterogeneous, although the overall conclusion across studies is consistent with the current study’s finding of a significant association between delirium and incident dementia. A recent large hospital administrative data study also accounted for competing mortality risk,^7^ and reported observed hazard ratios for incident dementia comparable to those in the current study. However, that study did not include prospective bedside delirium assessments and used hospital ICD-10 codes, which is likely to underestimate the prevalence of delirium (poorly coded in administrative data) and dementia (often only recorded in primary care data).^24,25^

In a study of 821 critically ill patients, Pandharipande *et al*. found that 24% of people who experienced delirium had cognitive scores at 12-month follow-up similar to those of people with mild Alzheimer’s disease, with deficits even in younger patients without preexisting conditions.^15^ The current study is aligned with these findings, although it is conducted in an older general medical population with a higher multimorbidity burden, and suggests that relative dementia risk following delirium may even be higher in those without preexisting conditions. In a cohort of 1510 adults aged ≥70 years, Tsui *et al*. found that people with higher baseline cognition were less likely to develop delirium, but when delirium occurred, it was often more severe and associated with greater future cognitive decline.^26^

Aligned with studies in a recent scoping review, people with delirium had a higher number of LTC than those without delirium.^9,27^ Only one study in that review explored the relationship between multimorbidity (measured using the Charlson Comorbidity Index (CCI)) and cognitive decline after delirium, in a selected subgroup of 126 older elective surgical patients.^14^ That study found multimorbidity contributed to explained variation in cognitive decline, with smaller decline in people with a higher CCI. The authors reported this to be in a “counterintuitive direction”, attributing this to limitations of their multivariable modelling approach and emphasising the need for replication. The current study supports their unexpected finding in a much larger, unselected population, demonstrating higher relative dementia risk following delirium in people without multimorbidity.

Our mortality findings align with those of a large single-centre study in Oxford, England, where delirium screening was performed using the Confusion Assessment Method and post-discharge ICD-10 codes used to generate a Hospital Frailty Risk Score.^27^ That study found relatively higher mortality risk associated with delirium in younger people (65-74 years), with lower frailty risk, and without dementia. In another cohort study of 710 adults ≥70 years, delirium on admission was associated with significantly higher mortality, with the highest relative hazard in fitter patients.^28^ The current study extends these findings, using linked primary care and hospital data to capture a broader range of LTC and shows that short-term mortality risk after delirium is high across the full spectrum of baseline health, including in those without multimorbidity. Taken together, studies support an interpretation of delirium as a marker of acute illness severity in older adults, independent of baseline health.

Key study strengths include the large, unselected sample of emergency admissions, increasing generalisability. Delirium assessments were performed directly and prospectively across multiple hospitals using the 4AT, a sensitive and specific tool effectively implemented in routine care and recommended in UK national guidelines,^18,29^ and 88% of potentially eligible participants had at least one 4AT during the study period. Multiple data sources were used to improve sensitivity of LTC and dementia ascertainment, including primary care records, important for identifying mental and behavioural conditions.^24^ Dementia diagnoses were based on clinical diagnoses in primary care and hospital records, previously shown to have high positive predictive value in this region compared to expert adjudication.^30^

We acknowledge several limitations. LTC (including dementia) may not have been diagnosed or recorded prior to admission. We mitigated this by ensuring participants had at least one year of GP registration before study entry,^16^ and by including conditions recorded during the index admission. Furthermore, we have previously shown that combining primary care, hospital, and community prescribing records captures ∼80% of expected dementia cases in this population.^31^ We used an unweighted LTC count for multimorbidity, and weighted indices such as the CCI or Elixhauser may be better associated with mortality.^21,32,33^ However, a condition count is easily clinically operationalisable and facilitated inclusion of a much wider range of conditions, including psychiatric and behavioural. Adjustment for acute illness severity was not possible, as physiological observations were not captured in EHR during the study period. These are now routinely electronically recorded and will be available for future research.

### Implications for Clinical Practice and Research

Study findings demonstrate that delirium in patients without preexisting dementia is strongly associated with incident dementia across the spectrum of multimorbidity. Delirium assessment should be performed rigorously in all acutely hospitalised older adults regardless of baseline health, as highlighted in national UK guidelines,^18^ and the high 4AT completion rate across multiple centres confirms feasibility. Delirium appears to be an important prognostic marker for mortality and dementia in hospitalised older adults, independent of baseline health. This study emphasises the need for post-delirium follow-up, which could potentially be tailored to baseline health. For example, people without multimorbidity may benefit from cognitive follow-up to detect and manage emerging cognitive decline, while those with multimorbidity may require holistic, multidisciplinary care to address overall health needs in the face of high mortality.^34^

Future research should investigate why people without multimorbidity are at relatively higher dementia risk following delirium, considering factors such as the precipitating event, delirium severity, and subtype. Specific patient subgroups, such as those with subclinical cognitive decline or genetic predisposition (*eg* apolipoprotein E genotype) may be at particularly high risk. Mechanisms to explain observed associations between delirium and dementia are incompletely understood, but biomarker and animal model studies suggest delirium is associated with markers of acute brain injury and may cause direct brain damage via processes including neuroinflammation, neurotransmitter dysregulation and disrupted cerebral metabolism.^35-39^ One prospective cohort study found delirium was associated with elevated blood levels of neuronal injury biomarker neurofilament light (NfL), and higher NfL levels were associated with greater cognitive decline.^37^ The Vantaa 85+ study found that dementia following delirium may involve a pathology trajectory distinct from other types of dementia.^40^ Greater understanding of neurodegenerative mechanisms in delirium could inform targeted interventions to reduce subsequent dementia risk, particularly in those with better baseline health.

## Conclusion

This study found that delirium on admission in hospitalised older adults was strongly associated with incident dementia across the spectrum of multimorbidity. These findings challenge the view that the adverse outcomes of delirium merely reflect baseline vulnerability and demonstrate that delirium is an important prognostic marker independent of multimorbidity burden. There is need to investigate underlying mechanisms of acute neurodegeneration in delirium and to develop targeted therapies with the potential to reduce future dementia risk.

## Acknowledgements

This work uses data provided by patients and collected by the NHS as part of their care. It has been facilitated by the DataLoch service (reference: DL_2023_012). DataLoch enables access to de-identified extracts of health care data from the South-East Scotland region to approved applicants: dataloch.org.

We would like to thank members of the Advanced Care Research Centre Patient and Public Involvement Network for their insights, which helped inform the study’s primary outcomes and ensure the research addressed questions relevant to older adults and their caregivers. With particular thanks to PPIE network member SF for reviewing the study and providing her insights on the findings and how to share these with patients and the public.

## Data and code availability

Data may be accessed through DataLoch (dataloch.org) following successful application and approvals. Analysis scripts for this study are available upon reasonable request to the corresponding author, with appropriate approvals from DataLoch.

## Copyright/ license for publication

For the purpose of open access, the author has applied a CC BY public copyright licence to any Author Accepted Manuscript version arising from this submission.

## Contributor and guarantor information

RSP, AMJM, AA and BG conceived the study and, along with CM, designed the methodology. RSP conducted the analysis. RSP, BG and AMJM produced the initial draft, and all authors including EWE, ELS and DHJD were involved in manuscript writing and editing. All authors critically reviewed and approved the final manuscript for submission. The corresponding author attests that all listed authors meet authorship criteria and that no others meeting the criteria have been omitted.

The lead author (the manuscript’s guarantor), Dr Rose S Penfold, accepts full responsibility for the work and/or the conduct of the study, had access to the data, and controlled the decision to publish. The lead author affirms that the manuscript is an honest, accurate, and transparent account of the study being reported; that no important aspects of the study have been omitted; and that any discrepancies from the study as originally planned have been explained.

