## Supplementary Materials for "Delirium is associated with incident dementia across the multimorbidity spectrum: a population-based cohort study"

**Supplementary Material**

**Table of Contents**

| **Supplementary Table 1**: Median number of days from admission to study end date in the delirium and no delirium groups | Page 2 |
| --- | --- |
| **Supplementary Table 2:** Read version 2 and ICD-10 code lists for dementia. | Page 3-5 |
| **Supplementary Table 3:** Long-term conditions in DataLoch primary care and hospital discharge data in relation to those included by “Measuring multimorbidity in research: a Delphi consensus study” (Ho *et al.,* 2022) | Page 6-10 |
| **Supplementary Table 4**: Frequency table showing number of admissions per individual during the study period | Page 11 |
| **Supplementary Table 5**: Characteristics of individuals excluded from the study with pre-existing dementia or no recorded 4AT score during the study period | Page 12 |
| **Supplementary Table 6**: Rates of death and new dementia diagnosis: overall, and stratified by delirium status and number of long-term conditions | Page 13 |
| **Supplementary Table 7:** At risk table, and number of people dead and diagnosed with dementia at each timepoint. | Page 14 |
| **Supplementary Table 8**: Sensitivity analysis, excluding patients who died and/or had a new dementia diagnosis in the six months following hospital discharge | Page 15 |
| **Supplementary Figure 1**: Study flowchart: inclusion, exclusion and creation of study cohort. | Page 16 |
| **Supplementary Figure 2**: Frequency distribution of number of long-term conditions in the study cohort | Page 17 |

**Supplementary Table 1:** Median number of days from date of index admission until study end date (23 October 2023) in the delirium and no delirium groups.

|  | Q1 | Median | Q3 |
| --- | --- | --- | --- |
| Delirium group | 1593 | 1892 | 2149 |
| No delirium group | 1617 | 1911 | 2169 |

**Supplementary Table 2:** Read version 2 and ICD-10 code lists for dementia (used for exclusion of patients with pre-existing dementia and identification of new dementia diagnoses).

Source: HDR UK Phenotype library, available at <https://phenotypes.healthdatagateway.org/phenotypes/PH148/version/296/detail/>

| **code** | **description** | **coding_system** |
| --- | --- | --- |
| **Eu01000** | [X]Vascular dementia of acute onset | Read codes v2 |
| **1461.00** | H/O: dementia | Read codes v2 |
| **E00y.11** | Presbyophrenic psychosis | Read codes v2 |
| **F110.00** | Alzheimer's disease | Read codes v2 |
| **E001.00** | Presenile dementia | Read codes v2 |
| **F110100** | Alzheimer's disease with late onset | Read codes v2 |
| **Eu01y00** | [X]Other vascular dementia | Read codes v2 |
| **Eu00000** | [X]Dementia in Alzheimer's disease with early onset | Read codes v2 |
| **E00y.00** | Other senile and presenile organic psychoses | Read codes v2 |
| **E00..00** | Senile and presenile organic psychotic conditions | Read codes v2 |
| **E001z00** | Presenile dementia NOS | Read codes v2 |
| **9Ou2.00** | Dementia monitoring second letter | Read codes v2 |
| **9hD0.00** | Excepted from dementia quality indicators: Patient unsuitabl | Read codes v2 |
| **Eu01111** | [X]Predominantly cortical dementia | Read codes v2 |
| **Eu00200** | [X]Dementia in Alzheimer's dis, atypical or mixed type | Read codes v2 |
| **Eu00z11** | [X]Alzheimer's dementia unspec | Read codes v2 |
| **E003.00** | Senile dementia with delirium | Read codes v2 |
| **E041.00** | Dementia in conditions EC | Read codes v2 |
| **E004.00** | Arteriosclerotic dementia | Read codes v2 |
| **E004100** | Arteriosclerotic dementia with delirium | Read codes v2 |
| **E002100** | Senile dementia with depression | Read codes v2 |
| **Eu01.11** | [X]Arteriosclerotic dementia | Read codes v2 |
| **E002z00** | Senile dementia with depressive or paranoid features NOS | Read codes v2 |
| **66h..00** | Dementia monitoring | Read codes v2 |
| **Eu02z11** | [X] Presenile dementia NOS | Read codes v2 |
| **Eu01.00** | [X]Vascular dementia | Read codes v2 |
| **E002.00** | Senile dementia with depressive or paranoid features | Read codes v2 |
| **8CMZ.00** | Dementia care plan | Read codes v2 |
| **Eu02z14** | [X] Senile dementia NOS | Read codes v2 |
| **E001200** | Presenile dementia with paranoia | Read codes v2 |
| **Eu01300** | [X]Mixed cortical and subcortical vascular dementia | Read codes v2 |
| **Eu01z00** | [X]Vascular dementia, unspecified | Read codes v2 |
| **Eu02z15** | [X] Senile psychosis NOS | Read codes v2 |
| **E004200** | Arteriosclerotic dementia with paranoia | Read codes v2 |
| **E004300** | Arteriosclerotic dementia with depression | Read codes v2 |
| **6AB..00** | Dementia annual review | Read codes v2 |
| **Eu00011** | [X]Presenile dementia,Alzheimer's type | Read codes v2 |
| **Eu00111** | [X]Alzheimer's disease type 1 | Read codes v2 |
| **E004.11** | Multi infarct dementia | Read codes v2 |
| **Eu02z12** | [X] Presenile psychosis NOS | Read codes v2 |
| **Eu00012** | [X]Primary degen dementia, Alzheimer's type, presenile onset | Read codes v2 |
| **Eu04100** | [X]Delirium superimposed on dementia | Read codes v2 |
| **E00..12** | Senile/presenile dementia | Read codes v2 |
| **E004z00** | Arteriosclerotic dementia NOS | Read codes v2 |
| **Eu02z13** | [X] Primary degenerative dementia NOS | Read codes v2 |
| **F110000** | Alzheimer's disease with early onset | Read codes v2 |
| **E002000** | Senile dementia with paranoia | Read codes v2 |
| **Eu00.00** | [X]Dementia in Alzheimer's disease | Read codes v2 |
| **Eu00113** | [X]Primary degen dementia of Alzheimer's type, senile onset | Read codes v2 |
| **Eu00z00** | [X]Dementia in Alzheimer's disease, unspecified | Read codes v2 |
| **9Ou4.00** | Dementia monitoring verbal invite | Read codes v2 |
| **9hD1.00** | Excepted from dementia quality indicators: Informed dissent | Read codes v2 |
| **9Ou5.00** | Dementia monitoring telephone invite | Read codes v2 |
| **9Ou3.00** | Dementia monitoring third letter | Read codes v2 |
| **9Ou1.00** | Dementia monitoring first letter | Read codes v2 |
| **Eu02z00** | [X] Unspecified dementia | Read codes v2 |
| **Eu00112** | [X]Senile dementia,Alzheimer's type | Read codes v2 |
| **Eu01100** | [X]Multi-infarct dementia | Read codes v2 |
| **9hD..00** | Exception reporting: dementia quality indicators | Read codes v2 |
| **E001000** | Uncomplicated presenile dementia | Read codes v2 |
| **E001300** | Presenile dementia with depression | Read codes v2 |
| **Eu01200** | [X]Subcortical vascular dementia | Read codes v2 |
| **ZS7C500** | Language disorder of dementia | Read codes v2 |
| **E00z.00** | Senile or presenile psychoses NOS | Read codes v2 |
| **E00..11** | Senile dementia | Read codes v2 |
| **Fyu3000** | [X]Other Alzheimer's disease | Read codes v2 |
| **Eu00100** | [X]Dementia in Alzheimer's disease with late onset | Read codes v2 |
| **E001100** | Presenile dementia with delirium | Read codes v2 |
| **E004000** | Uncomplicated arteriosclerotic dementia | Read codes v2 |
| **9Ou..00** | Dementia monitoring administration | Read codes v2 |
| **Eu02z16** | [X] Senile dementia, depressed or paranoid type | Read codes v2 |
| **Eu00013** | [X]Alzheimer's disease type 2 | Read codes v2 |
| **E000.00** | Uncomplicated senile dementia | Read codes v2 |
| **F05.1** | Delirium superimposed on dementia | ICD10 codes |
| **G30** | Alzheimer's disease | ICD10 codes |
| **F03** | Unspecified dementia | ICD10 codes |
| **F01** | Vascular dementia | ICD10 codes |
| **F00** | Dementia in Alzheimer's disease | ICD10 codes |

**Supplementary Table 3:** Long-term conditions in DataLoch primary care and hospital discharge data in relation to those included by “Measuring multimorbidity in research: a Delphi consensus study” (Ho *et al.*, 2022).

| **Ho et al. 2022 condition** | **Body system (based on ICD-10 chapters)** | **Always/ usually include** | **Present in DataLoch data** | **Data source: primary care and/or hospital discharge** | **List of DataLoch phenotypes for condition** |
| --- | --- | --- | --- | --- | --- |
| Heart failure | Cardiovascular disease | Always | Yes | Both | Heart failure |
| Chronic liver disease | Digestive disease | Always | Yes | Both | Alcoholic liver disease, Hepatic failure, Autoimmune liver disease |
| Diabetes | Metabolic and endocrine disease | Always | Yes | Both | Diabetes, Diabetic ophthalmic complications, Diabetic neurological complications |
| Parkinson's disease | Neurological disease | Always | Yes | Both | Parkinson's disease |
| End stage kidney disease | Urogenital disease | Always | Yes | Both | End stage renal disease |
| Coronary artery disease | Cardiovascular disease | Always | Yes | Both | Coronary heart disease not other specified, Myocardial infarction |
| Dementia | Mental and behavioural disorder | Always | Yes | Both | Dementia |
| Multiple sclerosis | Neurological disease | Always | Yes | Both | Multiple sclerosis |
| Stroke | Cardiovascular disease | Always | Yes | Both | Stroke NOS, Ischaemic stroke |
| Chronic kidney disease* | Urogenital disease | Always | Yes* | Biochemistry | Operationalised as per KDIGO criteria* |
| HIV/AIDS | Infectious disease | Always | Yes | Both | HIV |
| Metastatic cancers | Cancer | Always | Yes | Both | Secondary malignancy Liver and intrahepatic bile duct, Secondary malignancy Lymph Nodes, Secondary Malignancy Bone, Secondary Malignancy Bowel, Secondary Malignancy Lung, Secondary Malignancy retroperitoneum and peritoneum |
| Haematological cancers | Cancer | Always | Yes | Both | Multiple myeloma and malignant plasma cell neoplasms, Non-Hodgkin Lymphoma, Hodgkin Lymphoma, Leukaemia, Myelodysplastic syndromes |
| Solid organ cancers | Cancer | Always | Yes | Both | Primary malignancy biliary tract, Primary malignancy bladder, Primary malignancy bone and articular cartilage, Primary malignancy breast, Primary malignancy cervical, Primary malignancy colorectal and anus, Primary malignancy kidney and ureter, Primary malignancy liver, Primary malignancy lung and trachea, Primary malignancy mesothelioma, Primary malignany oesophageal, Primary malignancy oro-pharyngeal, Primary malignancy_other organs, Primary malignancy_other skin and subcutaneous tissue, Primary malignancy ovarian, Primary malignancy pancreatic, Primary malignancy prostate, Primary malignancy stomach, Primary malignancy testicular, Primary malignancy thyroid, Primary malignancy uterine |
| Cystic fibrosis | Metabolic and endocrine disease | Always | Yes | Both | Cystic Fibrosis |
| Epilepsy | Neurological disease | Always | Yes | Both | Epilepsy |
| Chronic obstructive pulmonary disease | Respiratory disease | Always | Yes | Both | Chronic obstructive pulmonary disease |
| Inflammatory bowel disease | Digestive disease | Always | Yes | Both | Ulcerative colitis, Chron's disease |
| Connective tissue disorder | Musculoskeletal disease | Always | Yes | Both | Giant Cell arteritis, Rheumatoid Arthritis, Lupus erythematosus, Psoriatic arthopathy, Systemic sclerosis, Ankylosing spondylitis |
| Paralysis | Neurological disease | Always | No | Neither | NA |
| Schizophrenia | Mental and behavioural disorder | Always | No | Neither | NA |
| Peripheral artery disease | Cardiovascular disease | Always | Yes | Both | Peripheral arterial disease |
| Asthma | Respiratory disease | Always | Yes | Both | Asthma |
| Addison's disease | Metabolic and endocrine disease | Usually | No | Neither | NA |
| Depression | Mental and behavioural disorder | Usually | Yes | Both | Depression |
| Heart valve disorders | Cardiovascular disease | Usually | Yes | Both | Multiple valve disease, Nonrheumatic aortic valve disorders, Nonrheumatic mitral valve disorders, Rheumatic valve dz |
| Bipolar disorder | Mental and behavioural disorder | Usually | Yes | Both | Bipolar affective disorder and mania |
| Melanoma | Cancer | Usually | Yes | Both | Primary malignancy Malignant Melanoma |
| Bronchiectasis | Respiratory disease | Usually | Yes | Both | Bronchiectasis |
| Osteoarthritis | Musculoskeletal disease | Usually | Yes | Both | Osteoarthritis (exc. Spine) |
| Chronic pancreatic disease | Digestive disease | Usually | Yes | Both | Pancreatitis |
| Arrhythmia | Cardiovascular disease | Usually | Yes | Both | Atrial fibrillation, Ventricular tachycardia |
| Thyroid disorders | Metabolic and endocrine disease | Usually | Yes | Both | Hypo or hyperthyroidism |
| Venous thromboembolic disease | Cardiovascular disease | Usually | Yes | Both | Venous thromboembolic disease (exc PE), Pulmonary embolism |
| Drug or alcohol misuse | Mental and behavioural disorder | Usually | Yes | Both | Alcohol problems, Other psychoactive substance misuse |
| Anaemia (including pernicious anaemia, sickle cell anaemia) | Haematological disorder | Usually | Yes | Both | Vit B12 deficiency anaemia, Other anaemias, Iron deficiency anaemia, Folate deficiency anaemias, Aplastic anaemias |
| Chronic Lyme disease | Infectious disease | Usually | No | Neither | NA |
| Transient ischaemic attack | Neurological disease | Usually | Yes | Both | Transient ischaemic attack |
| Eating disorders | Mental and behavioural disorder | Usually | Yes | Both | Anorexia and bulimia nervosa |
| Vision impairment that cannot be corrected | Eye disease | Usually | Yes | Both | Visual impairment and blindness |
| Long term musculoskeletal problems due to injury | Musculoskeletal disease | Usually | No | None | NA |
| Tuberculosis | Infectious disease | Usually | Yes | Both | Tuberculosis |
| Endometriosis | Urogenital disease | Usually | Yes | Both | Endometriosis |
| Chronic primary pain | Neurological disease | Usually | No | Neither | NA |
| Hearing impairment that cannot be corrected | Ear disease | Usually | Yes | Both | Hearing loss |
| Peptic ulcer | Digestive disease | Usually | Yes | Both | Peptic ulcer disease, Oesophageal ulcer and oesophagitis |
| Post-traumatic stress disorder | Mental and behavioural disorder | Usually | No | Neither | NA |
| Post-acute COVID-19 | Infectious disease | Usually | Not applicable to study period | Neither | NA |
| Benign cerebral tumours that can cause disability** | Cancer | Usually | No | Neither | NA |
| Peripheral neuropathy | Neurological disease | Usually | Yes | Both | Peripheral neuropathies (excluding cranial nerves and carpal tunnel syndromes) |
| Hypertension (treated and untreated) | Cardiovascular disease | Usually | Yes | Both | Hypertension |
| Congenital disease & chromosomal abnormalities | Congenital disease | Usually | No | None | NA |
| Chronic urinary tract infection*** | Urogenital disease | Usually | No | Neither | NA |
| Aneurysm | Cardiovascular disease | Usually | Yes | Both | Abdominal aortic aneurysm |
| Meniere's disease | Ear disease | Usually | Yes | Both | Meniere disease |
| Osteoporosis | Musculoskeletal disease | Usually | Yes | Both | Osteoporosis |
| Autism | Mental and behavioural disorder | Usually | Yes | Both | Autism and Asperger's syndrome |
| Anxiety | Mental and behavioural disorder | Usually | Yes | Both | Anxiety |
| Gout | Musculoskeletal disease | Usually | Yes | Both | Gout |

*Chronic Kidney Disease (CKD) phenotype was operationalised using KDIGO criteria (KDIGO CKD Work Group, 2024), specifically requiring that an individual’s most recent eGFR was <60 mL·min^−1^/1.73 m^2^ and at least one value obtained >90 days prior was also <60 mL·min^−1^/1.73 m^2^

**Not possible to determine degree of disability from hospital/ GP data. Inclusion of all relevant codes would significantly overestimate prevalence, therefore excluded

***Not possible to determine chronicity using hospital/ GP data. Inclusion of all UTI codes would significantly overestimate prevalence, therefore excluded

**Supplementary Table 4:** Frequency table, showing number of admissions per unique individual during study period 1 April 2017-1 April 2020.

| **No of admissions** | **Frequency** |
| --- | --- |
| 1 | 19301 |
| 2 | 6709 |
| 3 | 2743 |
| 4 | 1363 |
| 5+ | 1718 |

**Supplementary Table 5:** Characteristics of 8276 individuals excluded from analysis: 5065 with a recorded dementia diagnosis before or during index admission, and a further 3211 without a recorded 4AT during the study period.

|  | **All patients with dementia**  N = 5065 | **Dementia, no delirium**  **4AT 0-3**  N=1606 | **Dementia, delirium**  **4AT ≥ 4**  N=3099 | **Dementia, no recorded 4AT score**  N = 360 | **No dementia, no recorded 4AT score**  N= 3211 |
| --- | --- | --- | --- | --- | --- |
| **Age in years**  mean (SD) | 84.4 (6.9) | 83.9 (6.9) | 84.6 (6.9) | 84.0 (7.2) | 77.1 (7.8) |
| **Sex: Female** N(%) | 3047 (60.2) | 986 (61.4) | 1850 (59.7) | 211 (58.6) | 1492 (53.5) |
| **Ethnicity**  White  Other* | 4672 (96.0)  195 (4.0) | 1490 (96.2)  59 (3.8) | 2850 (95.9)  123 (4.1) | 332 (96.2)  13 (3.8) | 2841 (93.9)  184 (6.1) |
| **SIMD Quintile** N(%)  1 (most deprived)  2  3  4  5 (least deprived)  *21 missing* | 716 (14.2)  1089 (21.6)  748 (14.8)  1049 (20.8)  1451 (28.7) | 241 (15.0)  361 (22.5)  219 (13.7)  322 (20.1)  461 (28.7) | 437 (14.1)  655 (21.2)  472 (15.3)  641 (20.7)  885 (28.6) | 38 (10.6)  73 (20.3)  57 (15.9)  86 (24.0)  105 (29.2) | 438 (13.7)  757 (23.6)  552 (17.2)  547 (17.1)  908 (28.4) |
| **Number of LTC**  median (IQR) | 6 (4-8) | 6 (4-8) | 5 (4-7) | 5 (4-8) | 4 (2-6) |
| **LTC category**  No multimorbidity (0-1)  2-4  5-6  7+ | 168 (3.3)  1323 (26.1)  1399 (27.6)  2175 (42.9) | 54 (3.4)  382 (23.8)  454 (28.3)  716 (44.6) | 99 (3.2)  823 (26.6)  850 (27.4)  1327 (42.8) | 15 (4.2)  118 (32.8)  95 (26.4)  132 (36.7) | 619 (19.3)  1242 (38.7)  708 (22.2)  642 (20.0) |
| **Length of hospital stay (days)**  median (IQR) | 9 (3-28) | 8 (2-29) | 10 (3-31) | 2 (1-7) | 1 (0-4) |

SIMD: Scottish Index of Multiple Deprivation. LTC: long-term condition. SD: standard deviation. IQR: interquartile range

* Other categories combined into single group to avoid disclosure risks due to small numbers

**Supplementary Table 6:** Event rates of incident dementia and of death: overall, and by delirium status and number of long-term conditions.

| **Group** | **Number of people** | **Number of events** | **Person-years at risk** | **Event rate**  No of events/ 100 person-years (95% CI) |
| --- | --- | --- | --- | --- |
| **INCIDENT DEMENTIA***  Overall | 23558 | 2572 | 77281.4 | 3.3 (3.2-3.5) |
| No multimorbidity (0-1 LTC)  Delirium  No delirium  2-4 LTC  Delirium  No delirium  5-6 LTC  Delirium  No delirium  7+ LTC  Delirium  No delirium | 337  2700  1145  6686  1031  4828  1622  5209 | 59  133  243  681  199  478  240  539 | 978.1  11008.3  2693.9  25648.3  2021.8  16415.9  2741.8  15773.1 | 6.0 (4.6-7.8)  1.2 (1.0-1.4)  9.0 (7.9-10.2)  2.7 (2.5-2.9)  9.8 (8.5-11.3)  2.9 (2.7-3.2)  8.8 (7.7-9.9)  3.4 (3.1-3.7) |
| **MORTALITY**  Overall  No multimorbidity (0-1 LTC)  Delirium  No delirium  2-4 LTC  Delirium  No delirium  5-6 LTC  Delirium  No delirium  7+ LTC  Delirium  No delirium | 23558  337  2700  1145  6686  1031  4828  1622  5209 | 12488  183  775  805  2658  799  2553  1363  3352 | 121795.2  1684.7  13434.8  5918.6  34635.0  5293.4  25026.51  8319.9  27482.4 | 10.3 (10.1-10.4)  10.9 (9.3-12.6)  5.8 (5.4-6.2)  13.6 (12.7-14.6)  7.7 (7.4-8.0)  15.1 (14.1-16.2)  10.2 (9.8-10.6)  16.4 (15.5-17.3)  12.2 (11.8-12.6) |

LTC: long-term condition. CI: confidence interval.

*accounting for competing mortality risk

**Supplementary Table 7:** At risk table, and number of people (i) dead and (ii) diagnosed with dementia at 90 days, one year, two years and three years.

|  | **0 days** | **90 days** | | | | **365 days** | | | | **730 days** | | | | **1095 days** | | |
| --- | --- | --- | --- | --- | --- | --- | --- | --- | --- | --- | --- | --- | --- | --- | --- | --- |
|  | At risk | At risk | Death | Dementia  diagnosis | At risk | | Death | Dementia  diagnosis | At risk | | Death | Dementia  diagnosis | At risk | | Death | Dementia  diagnosis |
| Delirium, 7+ LTC | 1622 | 989 | 593 | 44 | 650 | | 838 | 137 | 473 | | 965 | 184 | 342 | | 1065 | 215 |
| Delirium, 5-6 LTC | 1031 | 617 | 370 | 44 | 437 | | 484 | 111 | 314 | | 569 | 148 | 243 | | 614 | 174 |
| Delirium, 2-4 LTC | 1145 | 756 | 342 | 47 | 547 | | 462 | 136 | 417 | | 544 | 186 | 337 | | 592 | 216 |
| Delirium, 0-1 LTC | 337 | 242 | 85 | 10 | 199 | | 107 | 32 | 169 | | 127 | 41 | 151 | | 139 | 47 |
| No delirium, 7+ LTC | 5209 | 4466 | 709 | 40 | 3681 | | 1359 | 171 | 3081 | | 1857 | 273 | 2538 | | 2300 | 374 |
| No delirium, 5-6 LTC | 4828 | 4212 | 580 | 40 | 3641 | | 1048 | 139 | 3185 | | 1405 | 239 | 2772 | | 1731 | 326 |
| No delirium, 2-4 LTC | 6686 | 6001 | 648 | 42 | 5419 | | 1096 | 172 | 4952 | | 1441 | 295 | 4476 | | 1779 | 432 |
| No delirium, 0-1 LTC | 2700 | 2515 | 175 | 10 | 2353 | | 310 | 38 | 2206 | | 426 | 68 | 2084 | | 526 | 90 |

**Supplementary Table 8:** Sensitivity analysis: unadjusted and adjusted subdistribution hazard ratios for incident dementia in hospitalised older adults with and without delirium, and by number of long-term conditions, excluding 5426 people who developed dementia (n= 617) and/or died (n= 4870) in the six months following hospital discharge.

|  | **Model 1**  Unadjusted  HR (95% CI) | **Model 2**  Adjusted for age, sex, SIMD  HR (95% CI) |
| --- | --- | --- |
| **INCIDENT DEMENTIA**^*^  No multimorbidity (0-1 LTC)  No delirium  Delirium  2-4 LTC  No delirium  Delirium  5-6 LTC  No delirium  Delirium  7+ LTC  No delirium  Delirium | Ref  3.99 (2.71-5.87)  2.26 (1.84-2.77)  5.90 (3.20-10.88)  2.21 (1.79-2.74)  5.48 (2.94-10.22)  2.40 (1.94-2.95)  4.79 (2.59-8.85) | Ref  3.65 (2.46-5.41)  1.53 (1.24-1.89)  3.46 (2.03-5.90)  1.44 (1.15-1.79)  3.07 (1.78-5.28)  1.51 (1.21-1.88)  2.72 (1.46-5.08) |

SIMD: Scottish Index of Multiple Deprivation. LTC: long-term condition. HR: hazard ratio.

*HR is the estimated subdistribution hazard ratio from a Fine-Gray hazard model accounting for competing mortality risk.

**Supplementary Figure 1:** Study flowchart: inclusion, exclusion and creation of study cohort.

EXCLUDED: DATA CHECKS

Recorded health activity after death: n= 25

Mismatch between date of birth and unique identifier: n=431

ELIGIBLE POPULATION

Registered with NHS Lothian GP for at least one year

n=305389 individuals

STUDY COHORT

Hospitalisation with one or more episodes of delirium 01/04/2017-01/04/2020: n=4135 unique individuals

Hospitalisation without delirium 01/04/2017-01/04/2020: n=19423 unique individuals

EXCLUDED

Dementia recorded before or during index admission: n= 5065 individuals

Hospitalisation with no 4AT recorded 01/04/2017-01/04/2020: n=3211 individuals

DELIRIUM STATUS ON ADMISSION

Delirium (4AT ≥4): n= 9250 admissions

No delirium (4AT 0-3): n= 39866 admissions

No 4AT recorded: n= 9174 admissions

EMERGENCY HOSPITALISATION

Emergency admission to medicine or orthopaedics from 01/04/2017 to 01/04/2020; over-65 at time of admission

n= 58290 admissions (31834 unique individuals)

**Supplementary Figure 2:** Histogram showing the distribution of number of long-term conditions in the study cohort.
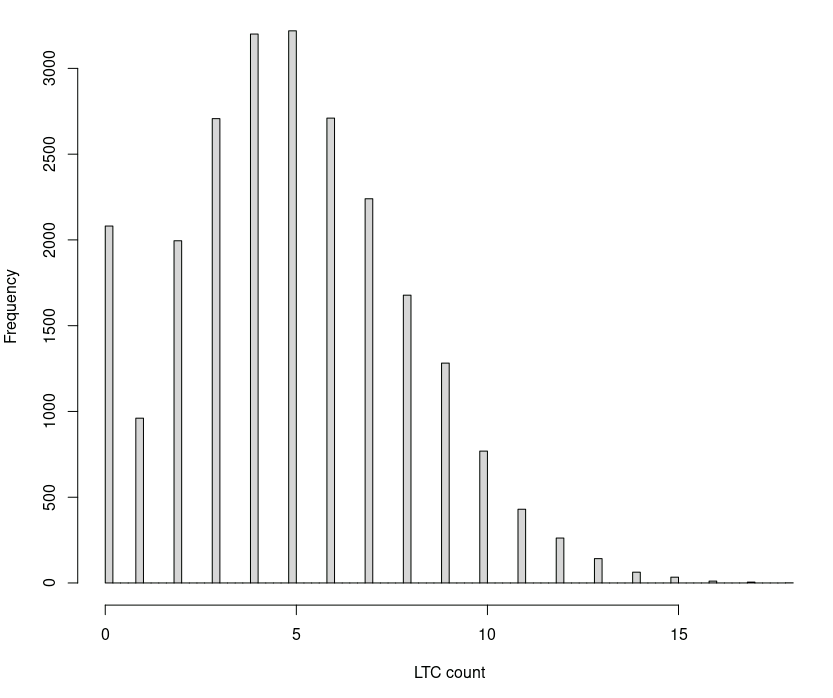
.

Median 5 (IQR 3-7); range 0-18
